# Compassionate Use of Opaganib For Patients with Severe COVID-19

**DOI:** 10.1101/2020.06.20.20099010

**Authors:** Ramzi Kurd, Eli Ben-Chetrit, Hani Karameh, Maskit Bar-Meir

**Affiliations:** Internal medicine, Shaare-Zedek Medical Center, Jerusalem, IL; Infectious diseases, Shaare-Zedek Medical Center, Jerusalem, IL; The Faculty of Medicine, Hebrew University, Jerusalem, IL

## Abstract

**Background:** Opaganib is a selective sphingosine-kinase (SK)-2 inhibitor with anti-inflammatory and anti-viral properties.

**Methods:** We provided opaganib on a compassionate-use basis to patients with severe COVID-19. Patients who required oxygen support via high-flow nasal cannula (HFNC) were offered the treatment. For comparison, we used a control group with same-sex, same-severity patients.

**Results:** Seven patients received at least one dose of opaganib since April 2, 2020. One patient, who received both hydroxychloroquine and azithromycin, developed diarrhea and all his medications were stopped. This was the only adverse effect possibly related to opaganib. A second patient was weaned of oxygen and discharged after receiving two doses of opaganib. Therefore, five patients were included in this analysis. Baseline characteristics were not significantly different between cases and controls. Patients treated with opaganib had significantly faster increase in lymphocyte count. All other clinical outcomes had a non-statistically significant trend in favor of the treatment group: median time to weaning from HFNC was 10 and 15 days in cases vs. controls (HR= 0.3, 95% CI: 0.07-1.7, p=0.2), time to ambient air was 13 vs.14.5 days (HR=0.4, 95% CI: 0.15-1.5), none of the cases required mechanical ventilation compared with 33% of controls.

**Conclusion:** In this small cohort of severe COVID-19 patients, opaganib was safe and well tolerated with improvement in both clinical and laboratory parameters in all treated patients. The efficacy of opaganib for COVID-19 infection should be further tested in randomized placebo-controlled trials.

## Introduction

Opaganib (RedHill Biopharma Ltd.) is a selective SK-2 inhibitor with anti-inflammatory and anti-viral properties. The stimulation of sphingolipid metabolism is a critical step in the mechanism of action of inflammatory cytokines. Sphingomyelin is recognized as a structural component of cellular membranes, but also serves as the precursor for the potent bioactive lipid ceramide and the proinflammatory lipid sphingosine 1-phosphate (S1P). Ceramide is produced by hydrolysis of sphingomyelin in response to inflammatory stress, including tumor necrosis factor alpha (TNFa)^1, 2^, and is able to induce apoptosis in proliferating cells. Ceramide can be further hydrolyzed by ceramidase to produce sphingosine, which is then phosphorylated by SK to produce S1P. Increasing intracellular levels of S1P and depletion of ceramide levels promote cell proliferation and inhibit apoptosis. S1P has been shown to protect neutrophils as well as macrophages from apoptosis^3, 4^, and was also shown to exert several other important effects on cells that mediate immune functions. Platelets, monocytes, and mast cells secrete S1P upon activation, promoting inflammatory cascades at the site of tissue damage^5, 6^. Activation of SK is essential for the signaling responses to TNFa^2^. Moreover, S1P mimics the ability of TNFa to induce the expression of cyclooxygenase-2 (COX-2) and the synthesis of prostaglandin E2 (PGE2)^7^. S1P is also a mediator of Ca^2+^ influx during neutrophil activation by TNFa leading to the production of superoxide and other toxic radicals^8, 9^. Opaganib (ABC294640) is an orally available inhibitor of the enzyme sphingosine kinase-2 (SK-2). In rodent models of inflammatory bowel and Crohn’s disease as well as in arthritis models, opaganib was noted to have an anti-inflammatory activity^10-12^. Furthermore, genetic deletion of SK-2 in mice showed amelioration of *Pseudomonas aeruginosa* induced lung injury^13^.

Several studies demonstrated the anti-viral effects of inhibiting SK-2 activity. Impairment of SK-2 expression or function significantly inhibited Chikungunya virus infection^14^, decreased the viral titers of Influenza virus in an in vitro model system^15^ and demonstrated a significant inhibitory effect in a preliminary Ebola cell-based inhibition assay^1^.

To date, three clinical trials have been completed with opaganib, a phase 1 study in advanced solid tumor patients^16^ and a food effect study in healthy volunteers as well as a phase 1b/2 in patients with advanced multiple myeloma (unpublished). In addition, two more studies are currently in progress, a phase 2 study in patients with advanced prostate cancer (clinicaltrials.gov ID: NCT04207255) and a phase 2 study in cholangiocarcinoma patients (clinicaltrials.gov ID: NCT03377179).

These anti-inflammatory and antiviral properties of opaganib, together with a good safety profile, have driven the use of opaganib under an expanded access program (compassionate use) in seven Israeli COVID-19 patients with severe disease. Here we report our experience with this treatment and compare the patients’ outcomes to those of untreated COVID-19 patients with a similar disease severity.

## Methods

### Patients and Setting

Shaare-Zedek Medical Center (SZMC) is a tertiary academic hospital in Jerusalem, IL. On February 27^th^ the first case of COVID-19 was diagnosed in Israel, with a dramatic increase in the number of cases following a Jewish holiday in the middle of March. Many cases were diagnosed in the orthodox Jewish communities of Jerusalem, which the hospital serves.

RedHill Biopharma, Ltd. offered opaganib under compassionate use for the treatment of COVID-19 patients. Eligible patients were those hospitalized with COVID-19 confirmed by a reverse-transcriptase-polymerase-chain-reaction assay. The treatment was offered to patients with severe disease requiring oxygen support via high-flow nasal cannula (HFNC). Patients who were intubated, those who could not provide informed consent, debilitated patients, pregnant or nursing women, patients on warfarin, apixaban, argatroban or rivaroxaban, and patients with New York Heart Association Class III or IV cardiac disease, myocardial infarction within the past 6 months, unstable arrhythmia, or evidence of ischemia on ECG, were excluded.

Screening for eligible patients was performed daily by two of the authors (R.K, E.B-C). Eligible patients were approached by one of the investigators (M.BM) and were offered opaganib treatment. The informed consent process was performed through remote video-calls. In addition to signing the informed consent, patients were required to have acceptable liver and renal function tests. [Bilirubin ≤ 1.5 times upper limit of normal (ULN), aspartate aminotransferase (AST) and alanine aminotransferase (ALT) ≤ 2.5 × ULN, Serum creatinine ≤ 1.5 X ULN], as well as acceptable hematologic status (absolute neutrophil count ≥1000 cells/mm^3^, platelet count ≥75,000 /mm^3^, hemoglobin ≥ 9 g/dL).

### Compassionate treatment

Oral opaganib was delivered at a dose of 500 mg q12 hours for up to 14 days, or until patient discharge. In order to monitor the tolerability of the medication, the first 3 patients received 250 mg q12 hours on days 1-3 and increased to the full dose on day 4. Subsequent patients were initiated at 500 mg q12 hours.

Treatment could have been discontinued if the patient or the treating physician decided that it is not in the patient’s best interest to continue. Additional therapy was allowed as deemed necessary by the clinicians.

All interventions were performed at the discretion of the treating physician. No additional tests or interventions were required in the opaganib treated patients..

Data were collected through May 5,2020.

### Assessment and analysis

Data on patients’ adverse events, oxygen-support requirements and laboratory values were collected daily until patient discharge. For comparison, we collected clinical data from a control group. To minimize bias we included in the control group same-sex, same-severity patients (male patients requiring HFNC oxygen support), aged 35 to 80 years, who met the inclusion and exclusion criteria stated above. These patient may have been eligible for the compassionate treatment, but were not approached due to time and staff constraints.

We compared clinical relevant endpoints such as time to weaning from HFNC (e.g low-flow oxygen via nasal cannula-NC); time to ambient (room) air (RA), as well as rate of C-reactive protein (CRP) decrease, and rate of lymphocyte count increase. For that purpose CRP and lymphocyte values were recorded at day 1 (treatment or admission) and at interval of 2-4 days. Time was calculated in days from the positive nasal swab. Other clinical parameters such as time to negative swab or time to chest X-ray (CXR) improvement, were not practical, because many patients were discharged prior to these endpoints.

We also recorded length of hospital stay, intensive care unit (ICU) admissions, need for intubation and mechanical ventilation and extracorporeal membrane oxygenation (ECMO) use in both groups.

### Ethical considerations

Individual approval was granted for each patient treated with opaganib by the SZMC institutional review board (IRB) and the Israeli Ministry of Health. Written informed consent was obtained from each participant. For the control group, IRB approval was granted to collect de-identified data. RedHill Biopharma Ltd. provided the medication, but was not involved in the conduct of treatment or follow-up, nor in the data collection and analyses. The authors assume full responsibility for the integrity and completeness of the data and have prepared the manuscript draft.

### Statistical analysis

Univariate comparisons between the groups were performed with chi-square test for categorical variables and t-test or Mann-Whitney U-test for continuous variables, as appropriate. Time variables were compared with Cox proportional hazard regression, adjusted for age and background illnesses. CRP and lymphocyte changes were compared utilizing a repeated measures general linear model with a Bonferroni correction for multiple comparisons. All analyses were conducted by one of the authors (M.B-M) with SPSS software, version 25.0 (IBM).

## Results

In total, seven patients received at least one dose of opaganib since April 2, 2020. All patients received hydroxychloroquine (HCQ), however one stopped HCQ prior to opaganib treatment due to borderline Q-T interval in ECG. Three patients received azithromycin as well. One patient, who received both HCQ and azithromycin, developed diarrhea after two doses of opaganib, and the treating physicians decided to stop all his medications. A second patient who deemed to be in severe condition, was weaned to low flow oxygen within hours, and was discharged on RA after receiving two doses of opaganib. Therefore, five patients were included in this analysis. Overall, 3 received the full 14-day course of opaganib, and 2 patients received 11 and 7 days respectively, before being discharged. All patients who received opaganib were male (approximately two-thirds of severe patients in SZMC were male).

### Safety

Except for diarrhea in one patient, noted above, no other adverse events related to the medication were observed. This patient had also started HCQ and azithromycin within a day of opaganib and therefore all three medications were stopped.

One patient reported dysuria on day 12 of treatment, that deemed to be unrelated to opaganib and another patient had an episode of atrial fibrillation that resolved spontaneously and did not recur despite continuation of opaganib.

### Baseline characteristics

Table 1 shows the characteristics of patients who received opaganib vs. the control cohort. Patients and controls had similar median age and similar rates of diabetes, hypertension and obesity. The median duration of symptoms prior to admission did not differ substantially between the groups. Cases had somewhat higher baseline lymphocyte counts and lower D-dimer levels compared with controls. All patients were defined by the treating physicians at baseline as severe COVID-19. All severe covid-19 patients received HCQ, and the majority also received azithromycin. However, methylprednisolone was given to a third of controls and to none of the cases.

**Table 1.** Baseline characteristics of the patients^*^.

| Characteristic | Opaganib<br>(N=5) | Control<br>(N=18) | *p* |
| --- | --- | --- | --- |
| Median age (IQR*) , years | 58 (50-73.2) | 58.5(54.5-66) | 0.9 |
| Median duration of symptoms before admission (IQR) , days | 7 (6-8) | 6(2.5-8) | 0.5 |
| Coexisting conditions -no.(%)<br>Hypertension<br>Diabetes mellitus<br>Obesity | 2(40)<br>2(40)<br>2(40) | 6(33)<br>6(33)<br>8(44) | 1 |
| Baseline laboratory values,<br>median (IQR)<br>CRP, mg/dL<br>Lymphocytes, mm<sup>3</sup><br>Fibrinogen, mg/dL<br>D-dimer, ng/mL<br>Ferritin, ng/mL | 15.8(12.6-21.7)<br>1100(995-1300)<br>858(844-2746)<br>1208(531-2633)<br>1483(999-5180) | 18.6(9-26.7)<br>850(635-970)<br>741(676-828)<br>1578(1073-3734)<br>758(530-1109) | 0.6<br>0.09<br>0.04<br>0.07<br>0.3 |
| Concomitant medications given for COVID-19, N (%)<br>hydroxychloroquine<br>azithromycin<br>methylprednisolone | 5(100)<br>3(60)<br>0 | 18(100)<br>14(77)<br>6(33) | 1<br>0.5<br>0.2 |

\*All patients are male; IQR- interquartile range

### Clinical outcomes

Among opaganib treated patients, median time to weaning from HFNC (time to NC) was 10 days (IQR: 9-15.5) compared with 15 days (IQR:8-19) in the control group, however this difference was not statistically significant [Hazard ratio (HR)= 0.3, 95% CI: 0.07-1.7, p=0.2] (Figure 1). Time to RA was 13 days in the opaganib group (IQR: 10.5-19) vs. 14.5 days in the controls (IQR: 10-28); (HR=0.4, 95% CI: 0.15-1.5).

**Figure 1.**
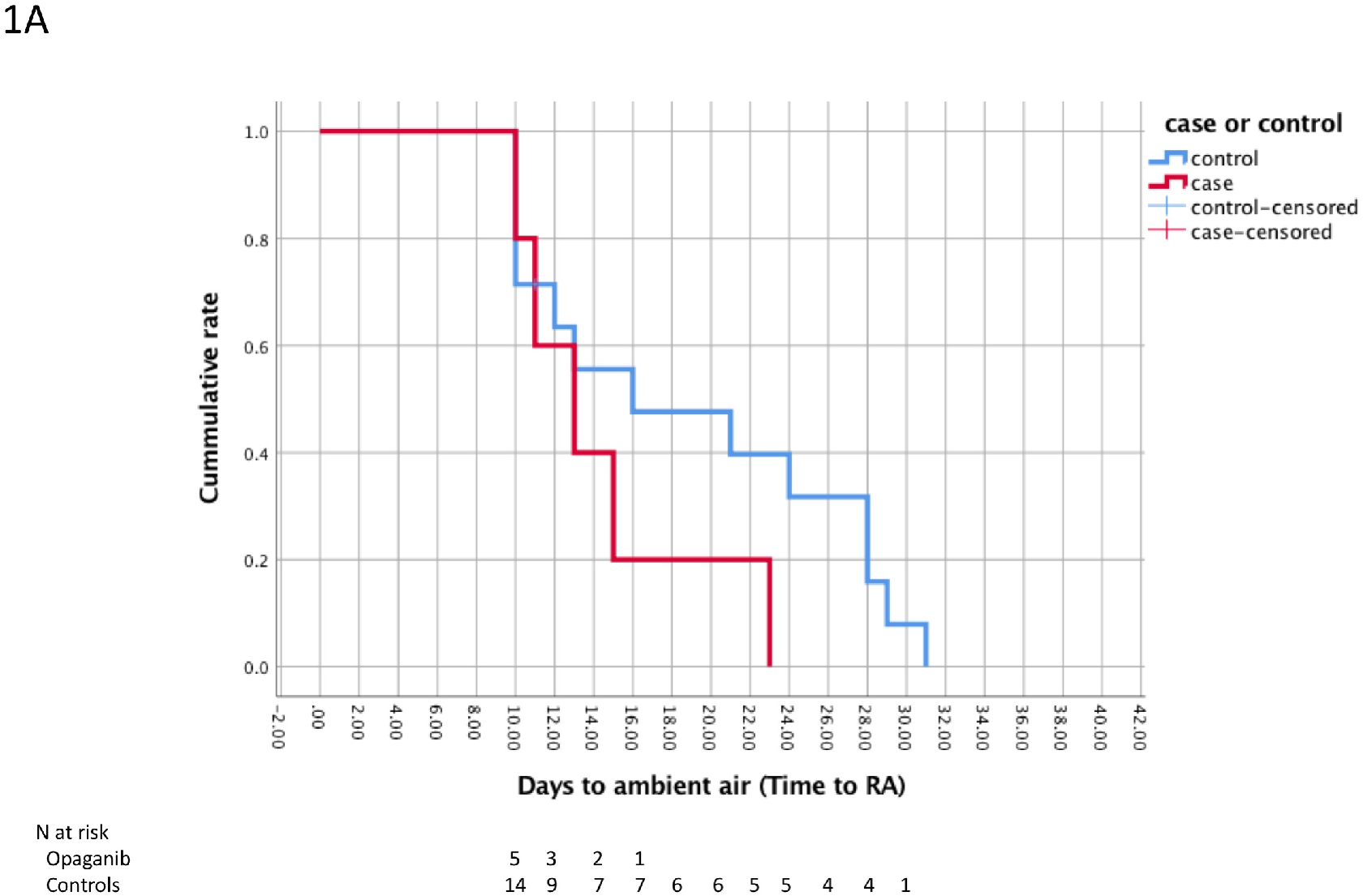
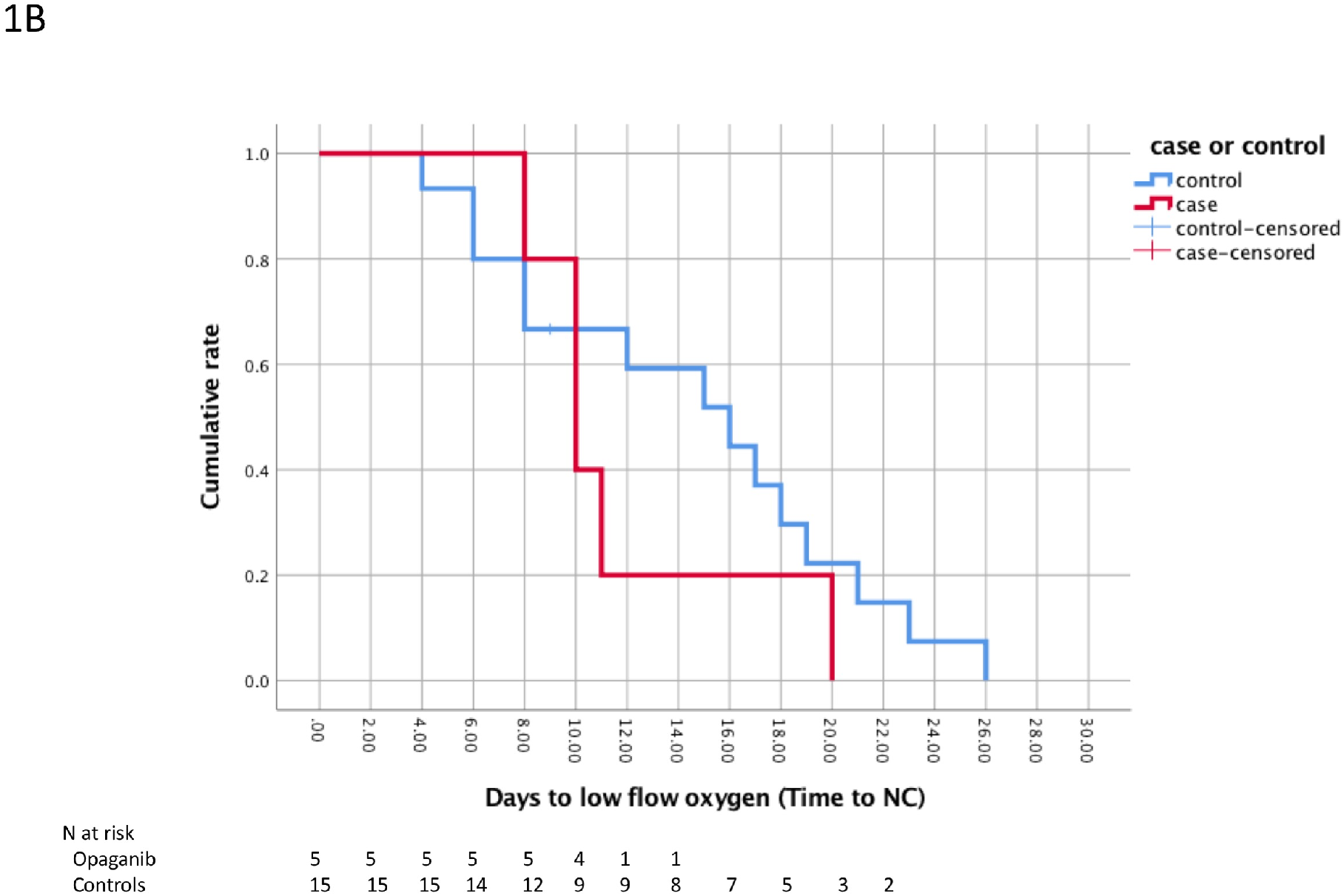
Cumulative rate of clinical improvement among cases vs. controls. 1A. Days from diagnosis to breathing ambient air; 1B. Days from diagnosis to low-flow oxygen support (weaning from high flow oxygen).

### Complications

All patients in the treatment group were discharged by the time of data collection, none have required mechanical ventilation or suffered complications. Among controls, 5 patients were not discharged by the time of data collection. Six patients in the control group (33%) required mechanical ventilation (p value compared with opaganib groups=0.13), 2 (11%) required ECMO, and one required tracheostomy. By the time of data collection, median length of stay was 18 days for the treatment group (IQR: 14-24.5) vs. 21.5 (IQR: 15.2-29.5) for controls. None of the patients in both groups had died by the time of data collection.

### Laboratory data

Lymphocyte counts increased significantly faster among the opaganib treated patients (p=0.001, Figure 2A). Of note, a third of the control patients received systemic steroids. In order to exclude bias related to the lymphocytopenic effect of steroids, the general linear model was performed again excluding these 6 patients, yielding similar results (p=0.013). The rate of CRP decrease in cases vs. controls did not reach statistical significance (p=0.08, Figure 2B).

**Figure 2.**
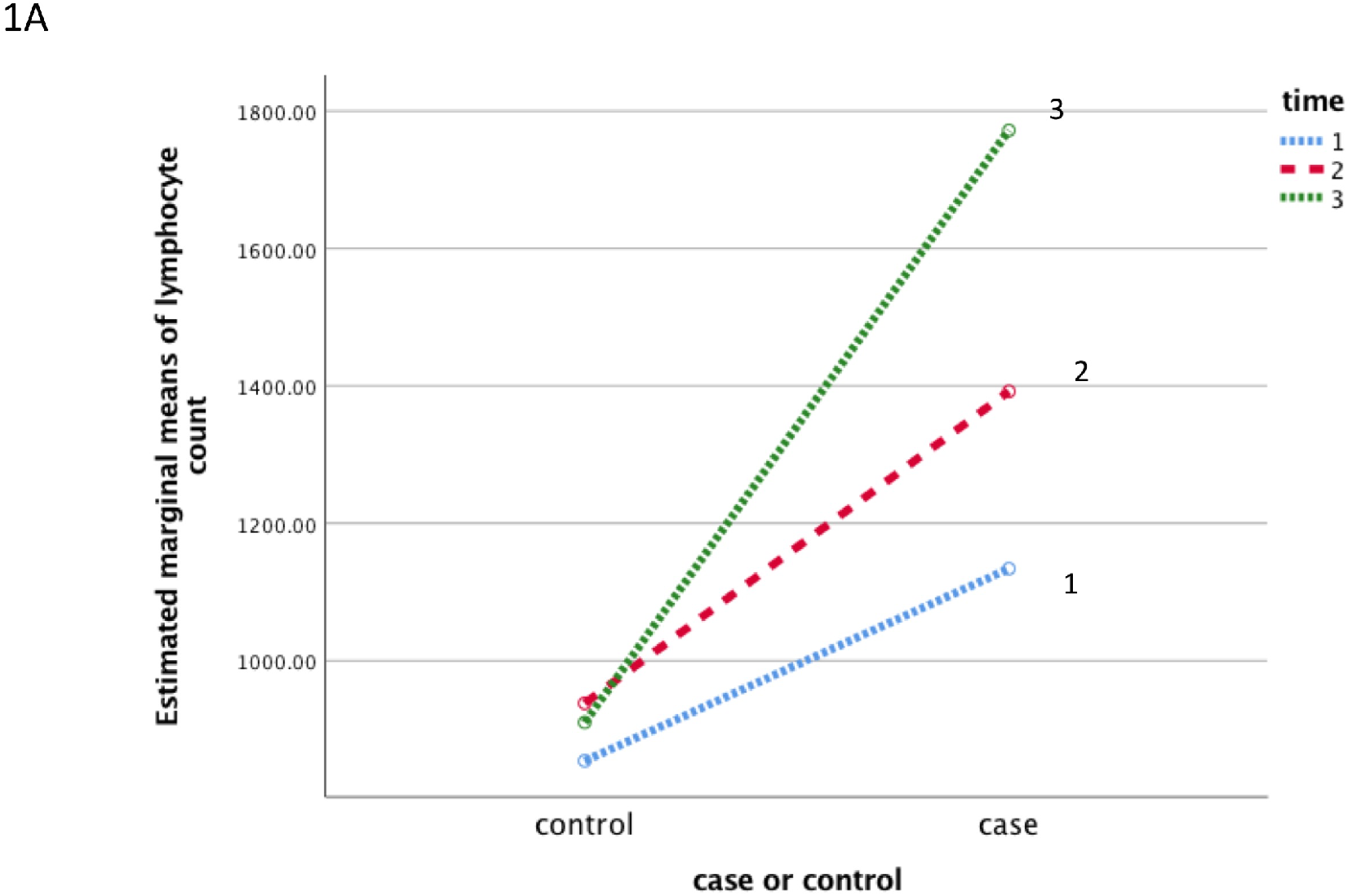
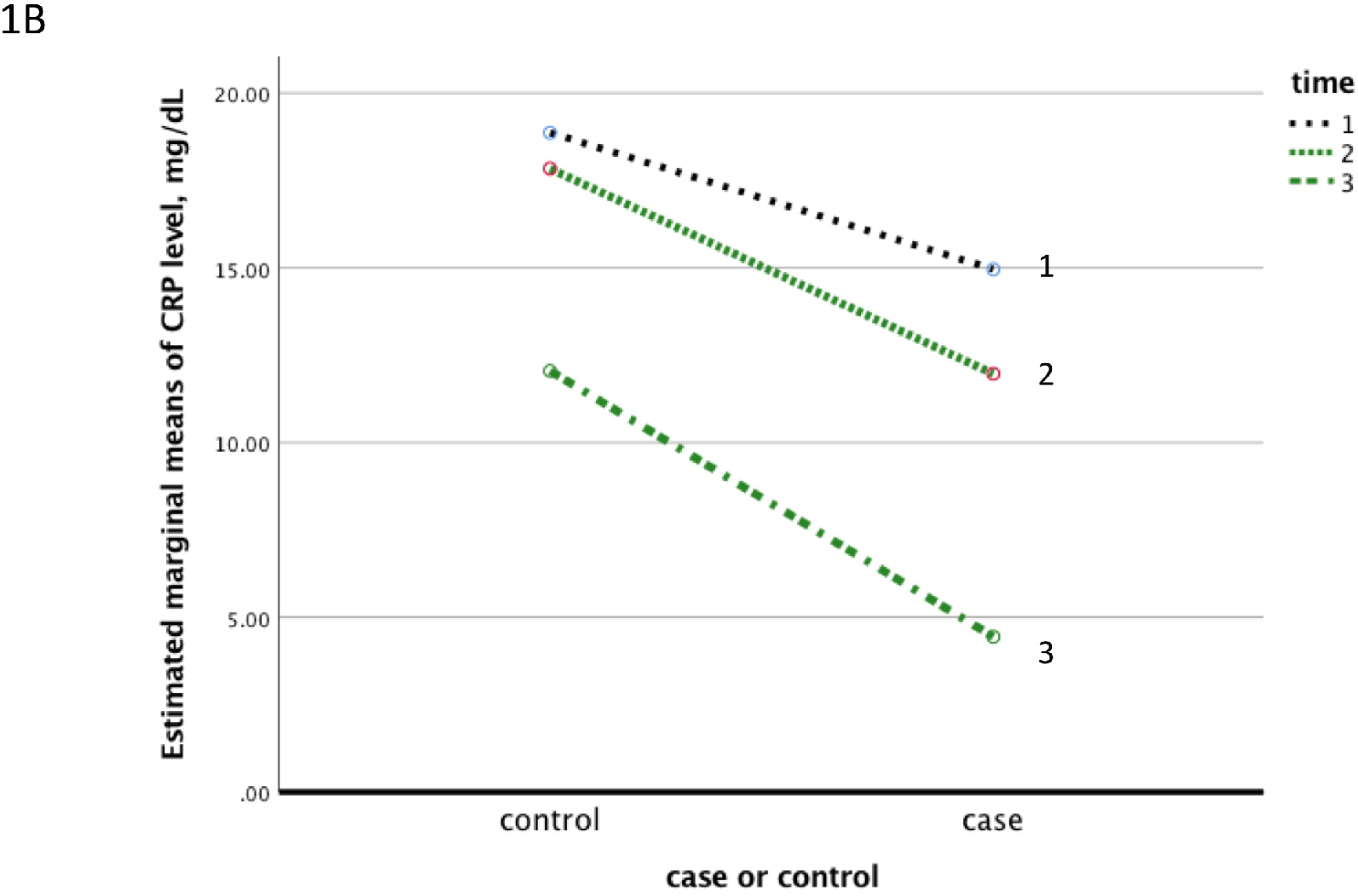
Results of analysis of variance by general linear model (GLM) repeated measures. 1A. lymphocyte count increased significantly faster in cases (right circles) vs. controls (left circles) over three time points (day of admission or day of treatment start and every 2-4 days); 2B-CRP increase over three time points in cases (right circles) vs. controls (left circles)

## Discussion

At the time of this study, no effective treatment was available for COVID-19 infection. Only at the end of this data collection, a preliminary report was published which showed some improvement in clinical outcomes of severe covid-19 patients treated with remdesivir^17^. Neither remdesivir nor other potential treatments such as convalescent plasma, were available for our patients. The anti-inflammatory and anti-viral properties of opaganib along with a good safety profile motivated us to offer it under compassionate use to severe covid-19 patients.

Although this report describes a very small cohort of patients, its preliminary results regarding the safety and efficacy of opaganib justify its further evaluation in larger cohorts, in a setting of a clinical trial.

Patients treated with opaganib had significantly faster increase in lymphocyte count. Low lymphocyte counts were associated with a more severe covid-19 disease ^18^ and with a more rapid deterioration^19^; therefore this faster increase may indicate better prognosis for the treatment group. Interestingly, the faster lymphocyte recovery among opaganib treated patients persisted even after excluding control patients who received systemic steroids. Although other clinical outcomes did not differ significantly between groups, the trends of time to improved oxygenation, rate of CRP decrease, as well as rates of mechanical ventilation, complications and discharge were all in favor of the treatment group.

Opaganib treatment was well tolerated. Only in one patient was treatment stopped, due to diarrhea, while being treated with HCQ and azithromycin in addition to opaganib. Of note, in a phase 1 advanced solid tumor trial with opaganib, neuropsychiatric effects, such as anxiety and hallucinations, were noted in up to 24% of patients. These side effects were diminished at doses < 750 mg q12 hours and if taken after a light meal. In our small cohort we did not observe such side effects. Neuropsychiatric side effects were described related to HCQ treatment in covid-19 patients ^20^, but also related to the infection itself ^21^. Therefore, a future clinical study of opaganib in a larger cohort of covid-19 patients will have to pay special attention to monitoring neuropsychiatric symptoms.

The results of the present report are preliminary and are limited by the small number of patients. However, in the absence of effective therapy for COVID-19, these data justify examining the effect of opaganib in the setting of a clinical trial, with larger numbers of patients and, potentially, with moderate disease severity.

## Data Availability

Data are not available at this point (pre-print)

1. R.Fathi, inventor Therapy for inhibition of single-stranded RNA virus replication U.S patent 10,543,222. 28 January, 2020.

## Notes

### Competing Interest Statement

The authors have declared no competing interest.

### Clinical Trial

NCT04435106

### Funding Statement

RedHill Biopharma Ltd. provided the medication, but did not fund the study, nor was it involved in the conduct of treatment, follow-up, data collection or analyses.

### Author Declarations

This work was approved by the Shaare-Zedek Helsinki committee #0123-20-SZMC and by the Israeli ministry of health

